# A monoclonal antibody-based immunoassay reinforces DOPA decarboxylase in cerebrospinal fluid as a diagnostic and prognostic biomarker for Parkinson’s disease

**DOI:** 10.1101/2025.02.26.25322938

**Authors:** Hubert Aviolat, Jennifer Mollon, Simone Giaisi, Stefan Barghorn, Roland G. Heym

## Abstract

**Background:** Novel supportive diagnostic and prognostic biomarkers for Parkinson’s disease (PD) are needed to enable its early diagnosis and inform clinical trials. Proteomic studies have identified cerebrospinal fluid (CSF) DOPA decarboxylase (DDC) as a promising biomarker candidate, but its role has not been well characterized. The aim of this study was to gain further insight into the potential of DDC as biomarker for PD.

**Methods:** We developed and validated a single molecule counting immunoassay for DDC quantification in CSF based on commercially available monoclonal antibodies. DDC levels were quantified in the Parkinson’s Progression Markers Initiative cohort including healthy controls (n=29), dopaminergic drug-naïve patients with PD (n=27) and with scans without evidence for dopaminergic deficit (SWEDD) (n=18). Their relationship with ioflupane-[123I]-single-photon emission computed tomography-based dopamine transporter (DaT-SPECT) imaging was analyzed. The prognostic potential of CSF DDC was evaluated by assessing the relationship between baseline DDC levels and yearly changes of Movement Disorder Society Unified Parkinson’s Disease Rating Scale (MDS-UPDRS) scores. CSF DDC levels were also quantified three years after the diagnosis, and their relationship with the L-DOPA equivalent daily dose (LEDD) was investigated. Finally, absolute DDC levels determined by our assay were correlated with relative concentrations obtained from Olink technology.

**Results:** Our DDC assay detected elevated levels in CSF from dopaminergic drug-naïve PD patients and discriminated them from untreated SWEDD and control with high sensitivity and specificity. There was an inverse correlation between baseline DDC levels and DaT-SPECT striatal binding ratios (SBR) from the putamen and caudate nucleus. Baseline CSF DDC levels demonstrated prognostic potential for MDS-UPDRS total change five to eight years after the diagnosis. DDC levels were further increased at the three-year follow-up visit in PD patients and were positively correlated with the LEDD. Finally, there was a strong correlation between relative CSF DDC levels determined with the Olink assay and absolute DDC levels determined with our assay.

**Conclusions:** Our monoclonal antibody-based assay for DDC quantification provided further insight into the potential of DDC in CSF to serve as a diagnostic and prognostic biomarker for PD. The unchanged levels in SWEDD patients and the inverse correlation with DaT-SPECT SBR suggest that DDC levels in CSF are connected to dopaminergic deficit.

## Background

Parkinson’s disease (PD) is the second most prevalent neurodegenerative disorder, and its incidence and public health burden continue to rise [1,2]. The diagnosis of PD remains based on clinical assessment. Ancillary tests, such as imaging or genetic tests, are generally reserved for individuals with an atypical clinical presentation [3,4]. Tissue and fluid biomarkers for PD are also being investigated, with α-synuclein seed amplification assays (SAAs) in cerebrospinal fluid (CSF) and skin samples demonstrating high performance in differentiating patients with PD from healthy controls (Ctrl) and detecting individuals in the prodromal phase [5–8]. However, the timely diagnosis of PD remains challenging, highlighting the need to develop further supportive diagnostic biomarkers [9]. Moreover, striking heterogeneity has been observed among patients with PD with regards to their disease etiology, clinical presentation, progression rate, and treatment response, emphasizing the significance of identifying prognostic biomarkers and implementing a precision medicine approach [3,9].

DOPA decarboxylase (DDC) is implicated in dopamine biosynthesis [10,11]. It has recently emerged as a potential biomarker for PD, demonstrating elevated levels in CSF of patients with PD, including those who are dopaminergic drug-naïve or in the prodromal phase [12–17]. In addition, increased DDC levels have been detected in CSF from patients with atypical Parkinsonian syndromes, including dementia with Lewy bodies [13,15,16]. Potential reasons for the elevation of CSF DDC have been investigated and an association with both dopaminergic treatment and dopamine transporter imaging have been reported [12,13,17]. However, the interpretation of these findings is complicated by the interconnected nature of dopaminergic deficit, dopaminergic treatment, and disease severity. Therefore, further studies are needed to better understand the reasons for increased CSF DDC levels, its changes with disease progression, and its ability to serve as a prognostic biomarker.

Most previous publications have relied on proprietary proximity extension assay technology by Olink for DDC detection [12–18]. A broadly accessible, sensitive, and quantitative immunoassay for CSF DDC assessment is needed to confirm the previous findings and accelerate research on DDC as a biofluidic biomarker.

Here, we describe the development and validation of a sensitive immunoassay for absolute DDC quantification in CSF using commercially available monoclonal antibodies. Using this assay, we quantified CSF DDC levels in healthy controls, patients with PD, and patients with scans without evidence for dopaminergic deficit (SWEDD) from the Parkinson’s Progression Markers Initiative (PPMI) cohort at baseline and 3-year follow-up to gain further insight into the potential of DDC as a diagnostic and prognostic biomarker for PD.

## Methods

### Study participants

#### Assay validation cohort

Assay validation was performed in a cohort including CSF samples from 10 patients with PD and 10 Ctrl. Their demographic characteristics are presented in Supplementary Table S1. The samples were obtained from PrecisionMed, LLC (Carlsbad, CA, USA). CSF samples were diluted 1:10, and all conditions were tested in duplicates.

#### Parkinson’s Progression Markers Initiative (PPMI) cohort

CSF samples from individuals from the PPMI cohort were included in the study for DDC level quantification [19]. Baseline and three-year follow-up CSF samples from 27 dopaminergic drug-naïve patients with PD, 18 dopaminergic drug-naïve patients with SWEDD and, 29 Ctrl were analyzed.

Several other measures and characteristics that were available for the study participants from the PPMI cohort were also used in the study. Ioflupane [123I]-single-photon emission computed tomography (SPECT)-based dopamine transporter imaging (DaT-SPECT) striatal binding ratio (SBR) from the study participants were available at baseline [4]. The L-DOPA equivalent daily dose (LEDD) was considered at the three-year follow-up visit [20–22]. Yearly Movement Disorder Society Unified Parkinson’s Disease Rating Scale (MDS-UPDRS) score changes for years 1 through 8 were also incorporated in the study [23]. For the MDS-UPDRS score changes, in addition to the total score OFF, the subscores for Part I, Non-Motor Aspects of Experiences of Daily Living; Part II, Motor Aspects of Experiences of Daily Living and Part III OFF, Motor Examination were also considered. A previous analysis of MDS-UPDRS subscores in the PPMI cohort revealed a continual measured progression of motor and non-motor symptoms during the initial five years following PD diagnosis [24].

### Identification and screening of antibodies for a DDC immunoassay on the Mesoscale Discovery (MSD) platform

We searched the literature for commercially available anti-DDC antibodies. Seven monoclonal antibodies targeting different DDC epitopes and one polyclonal antibody raised against the full-length DDC protein were selected for further analysis (Supplementary Table S2). The antibody buffer was exchanged to phosphate-buffered saline (PBS) in all instances using desalting column (Thermo Fisher Scientific, 87764). Three recombinant DDC proteins from different expression systems (bacterial, insect, and human cells) were tested as calibrators (Supplementary Table S3). The antibodies and calibrators were quality controlled using NanoDrop and SDS-PAGE. Next, 70 µg and 15 µg of antibodies were used for biotin (Thermo Fisher Scientific, 90407) and for SULFO-TAG labeling (Meso Scale Discovery, R91AO), respectively. Calibrators were tested at 1 ng/mL. CSF samples diluted 1:5 and 1:20, plasma samples diluted 1:10 and 1:40, eight antibodies and, three standard proteins were analyzed in LowCross-Buffer MILD (Candor; 101 500). MSD GOLD 96-well Small Spot Streptavidin SECTOR plates were used (Meso Scale Discovery, L45SA). A 3.5 µg/mL capture antibody was incubated for 1 h at room temperature and 700 rpm in Diluent 100 (Meso Scale Discovery, R50AA). The samples were then incubated for 2 h at room temperature and 700 rpm. Next, a 0.75 µg/mL detection antibody was added and incubated for 1 h at room temperature and 700 rpm in LowCross-Buffer MILD. Finally, MSD GOLD Read Buffer B (Meso Scale Discovery, R60AM) was added. The signal-to-noise (S/N) ratio was used to screen for an optimal combination of a capture antibody, detection antibody, and calibrator protein.

### Development of an ultrasensitive single molecule counting (SMC) immunoassay

The antibodies, samples, and calibrator were diluted in LowCross Buffer® MILD (Candor; 101 500). Anti-human DDC mouse monoclonal 413911 antibody (R&D Systems; MAB3564) at 17.5 µg of antibody/mg of beads (Merck; 03-0077-02) was used as the capture antibody. Recombinant human DDC (rhDDC) protein (R&D Systems; 3564-DC-010) was employed as a calibrator. Capture antibody-coated beads (10 mg of beads per mL) were mixed at 1,250 rpm until all beads were resuspended and diluted 200-fold. Next, 100 µL of diluted beads and 100 µL of samples (human CSF samples diluted 1:10) or calibrators were added to each well of Axygen Assay Plates (Axygen; P-96-450V-C), which were incubated for 2 h at 25 °C with shaking (Jitterbug; setting Mix 5). The plates were washed with Erenna System buffer (Merck, 02-0111-03) using a Tecan Hydroflex washer. After aspiration, 20 μL of a filtered fluorescently labeled (Merck; 03-0076-02) anti-human DDC mouse monoclonal A17096A antibody (BioLegend; 869501) at 937.5 ng/mL, which served at the detection antibody, were added to each well and incubated for 1 h at 25 °C with shaking (Jitterburg, setting Mix 5). The plates were washed as previously. The beads were transferred to a fresh Axygen 96-well plate, into 200 μL of Erenna System buffer. After final aspiration, 9.5 μL of Elution Buffer B (Merck; 02-0297-00) were added to each well of the assay plate, and the plates were incubated for 20 min at 25 °C with shaking (Jitterburg, setting Mix 5). The eluate was magnetically separated from the beads, transferred into a 384-well plate (Thermo Fisher Scientific; 264573) and neutralized immediately with 9.5 µL/well of Buffer D (Merck, 02-0368-00). After heat-sealing (4titude; 4ti-0531), the plate was analyzed using the Erenna instrument

#### *In vitro* validation of the SMC DDC assay

The newly developed assay was analytically validated for its sensitivity, parallelism, spike-in recovery, and dilution linearity. In all instances, the validation criteria were ±30% for recovery and coefficient of variability (CV). To determine the assay’s limits of quantification, the rhDDC protein was tested in concentrations of 0.316–10,000 pg/mL. A five parameters logistic regression, weight by 1/Y^2^ was performed, and the lower limit of quantitation (LLOQ) and upper limit of quantitation (ULOQ) were determined (corresponding to the lowest and highest concentration of the calibration standards that fulfill validation criteria). To assess the parallelism of the assay, samples pooled from 5–6 individuals were tested. The spike-in recovery of rhDDC in human CSF was tested by measuring the recovered DDC concentrations for 0, 10, 100, and 1,000 pg/mL DDC calibrator protein spiked into biological matrix. Both the mean of four samples per spike concentration and 75% of individual samples per spike concentration had to match the recovery validation criterion. The dilution linearity of rhDDC in human CSF was assessed for concentrations of 0.06–2,000 pg/mL.

### DDC assay conditions for the PPMI cohort (Project 274)

All samples from the PPMI cohort were run on the same day on four plates with four quality control samples (CSF pools) on each plate. Because the CV of the quality control samples was low (<6%) (Supplementary Figure S1), no data correction was applied across the plates. Finally, the correlation of DDC levels quantified with the newly developed SMC assay and DDC levels determined with a proteomic Olink Explore assay (cardiometabolic panel) was explored in a subset of samples.

#### Analysis of the prognostic potential of DDC CSF levels in patients with PD

Baseline DDC levels in CSF from dopaminergic drug-naïve patients with PD were split at the median into low-DDC (13 patients) and high-DDC (14 patients) groups. Subsequently, changes in the MDS-UPDRS scores in the low-DDC and high-DDC groups over time as well as correlations between DDC levels and MDS-UPDRS scores were evaluated. A cutoff of 8 years ensured a minimum of 14 patients with available MDS-UPDRS scores and baseline DDC levels in both the low-DDC and high-DDC groups.

### Statistical analysis

Numerical data are presented as mean ± standard deviation (SD) or median with 95% confidence interval (CI). A receiver operating characteristic (ROC) curve was plotted, and the area under the curve (AUC) and the sensitivity and specificity of the DDC assay in the validation PPMI cohorts were calculated. For statistical group comparisons, methods used are thoroughly described in corresponding figure legends. Spearman’s rank correlation was used to investigate the correlation between two parameters.

## Results

### Development and validation of an ultrasensitive SMC assay for DDC quantification in CSF

Initially, we screened anti-DDC antibodies in an MSD assay format. However, for CSF samples, all combinations of potential capture and detection antibodies with acceptable signal to noise (S/N>2) ratios included at least one polyclonal antibody (Supplementary Figure S2). Therefore, we selected the most promising combination of monoclonal antibodies and calibrator and set out to develop a novel SMC assay. Analytical validation of the SMC DDC assay using rhDDC protein revealed an estimated LLOQ of 1 pg/mL and estimated ULOQ of 1,000 pg/mL (Figure 1a). For human CSF samples, parallelism was observed for 1:4–1:64 dilutions (Figure 1b). In the spike-in recovery analysis, the high and medium concentration of spiked DDC fell in the acceptance range (Figure 1c). For the low concentration of spiked DDC (10 pg/mL), the mean of four samples met the ±30% acceptance criterion, but only one out of the four individual samples did (Figure 1c). The dilution linearity of rhDDC in human CSF met the acceptance criteria (Figure 1d).

**Figure 1.**
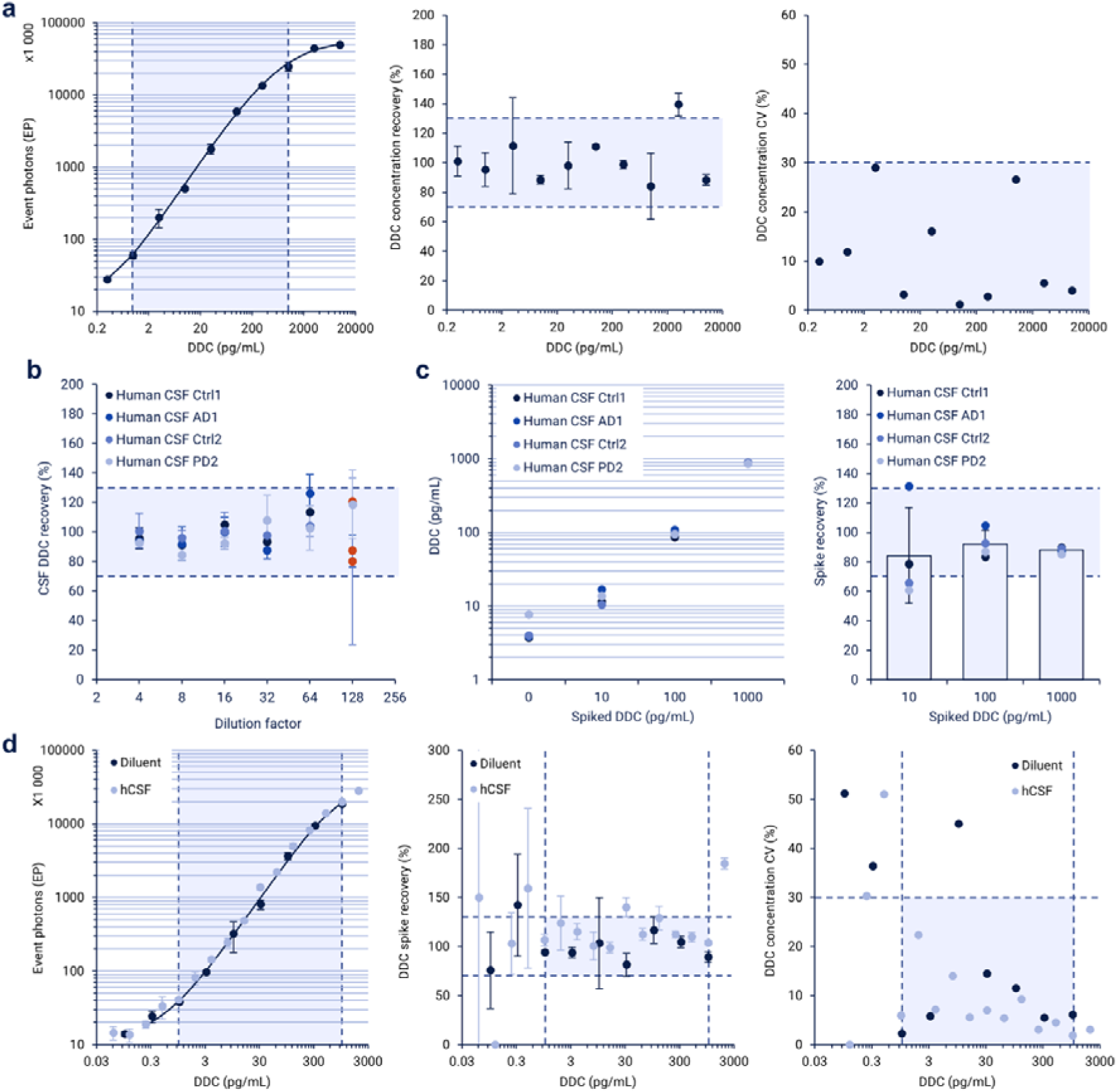
*In vitro* validation of the SMC DDC assay, including limits of quantification (**a**), parallelism (**b**), spike-in recovery (**c**), and dilution linearity (**d**). Mean ± SD of technical duplicates are plotted. Vertical dashed lines represent LLOQ and ULOQ. Horizontal dashes lines represent the +/-30% validation criterium.

We used our validated SMC assay to quantify DDC in CSF samples from a small pilot cohort of patients with PD (n = 10) and Ctrl (n = 10) (Supplementary Table S1). DDC levels were significantly higher in patients with PD than in the control group (p = 0.0001) (Supplementary Figure S3).

### The new DDC SMC assay has high discriminatory power for dopaminergic drug-naïve PD patients

Next, CSF DDC levels were quantified in a subset of the PPMI cohort, including Ctrl (n = 29) and dopaminergic drug-naïve patients with PD (n = 27) or with SWEDD (n = 18). The demographic baseline characteristics of the participants are presented in Table 1. At baseline, CSF DDC levels were higher in dopaminergic drug-naïve patients with PD than in Ctrl (1.62-fold; p<0.0001) and patients with SWEDD (1.64-fold; p<0.0001). No significant difference was detected between the CSF DDC levels of Ctrl and patients with SWEDD (p=0.5040) (Figure 2a). There were no significant age differences among the groups (Supplementary Figure S4a) as well as no significant correlation between CSF DDC levels and age (Supplementary Figure S4b). No significant differences in DDC levels were observed between males and females in each group (Supplementary Figure S4c). There was no significant correlation between CSF DDC levels and hemoglobin contamination levels (Supplementary Figure S4d).

**Table 1.** Demographic characteristics of the tested PPMI cohort at baseline. Ctrl: healthy control; PD: Parkinson’s disease; SWEDD: Scans without evidence for dopaminergic deficit

|  | Ctrl<br>(n = 29) | PD<br>(n = 27) | SWEDD<br>(n = 18) |
| --- | --- | --- | --- |
| **Age (years)** |  |  |  |
| Mean (SD) | 63.8 (10.8) | 61.7 (9.1) | 59.6 (10.6) |
| Range | 31.2-81.9 | 38.5-75.6 | 38.3-75.7 |
| **Sex (Number, %)** |  |  |  |
| Male | 18 (62.1) | 19 (70.4) | 12 (66.7) |
| Female | 11 (37.9) | 8 (29.6) | 6 (33.3) |

**Figure 2.**
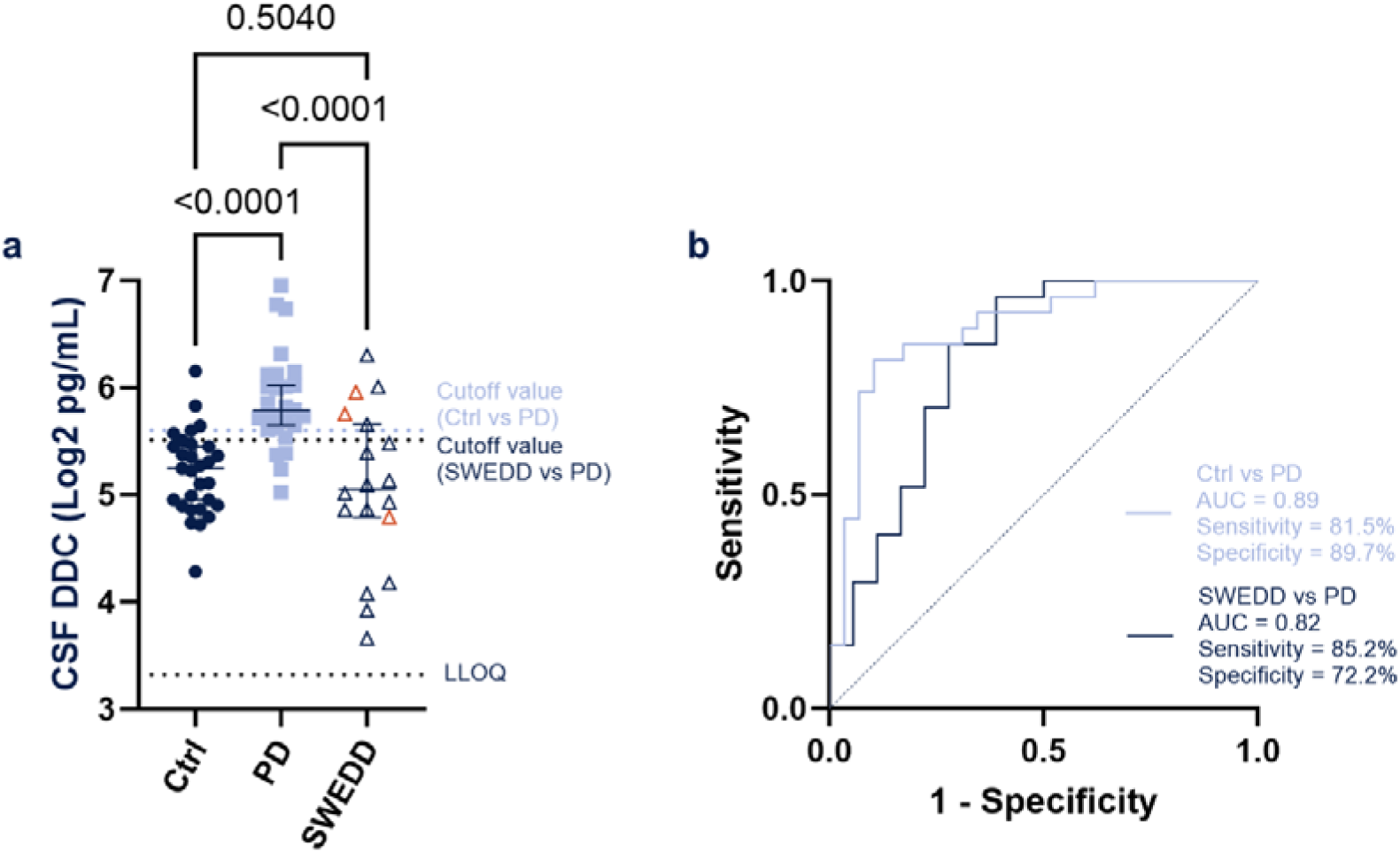
DDC levels in CSF of dopaminergic drug-naïve patients with PD and with scan without evidence of dopaminergic deficit SWEDD and Ctrl. **a**. Comparison of CSF DDC levels among the groups. For each individual, the mean value of technical duplicates is plotted. Horizontal bars correspond to median ± 95% CI. CSF SAA+ SWEDD are highlighted in red. P-values are from multiple linear regression analysis of log2 CSF DDC data, including sex and age as covariates. **b**. Receiver operating characteristic analysis.

The discriminatory performance of DDC for dopaminergic drug-naïve patients with PD was assessed using a ROC curve analysis. It revealed an AUC of 0.89 (p<0.0001) and 81.5% sensitivity and 89.7% specificity to discriminate dopaminergic drug-naïve patients with PD from Ctrl and an AUC of 0.82 (p = 0.0004) and 85.2% sensitivity and 72.2% specificity to discriminate dopaminergic drug-naïve patients with PD from patients with SWEDD (Figure 2b).

In addition, there was a strong correlation between CSF DDC levels determined with the Olink Explore assay in two different studies and the SMC assay developed in the current study (r = 0.76; p = 0.0002 and r = 0.71; p<0.0001) (Supplementary Figure S5).

### CSF DDC levels in patients with PD correlate with DaT-SPECT SBR

At baseline, CSF DDC levels showed a significant inverse correlation with DaT-SPECT SBR of the contralateral caudate and putamen (Spearman r = -0.51; p<0.0001 and r = -0.54; p<0.0001, respectively) when analyzing all dopaminergic drug-naïve disease groups (Figure 3). However, no significant correlation was observed when analyzing only the PD group (data not shown).

**Figure 3.**
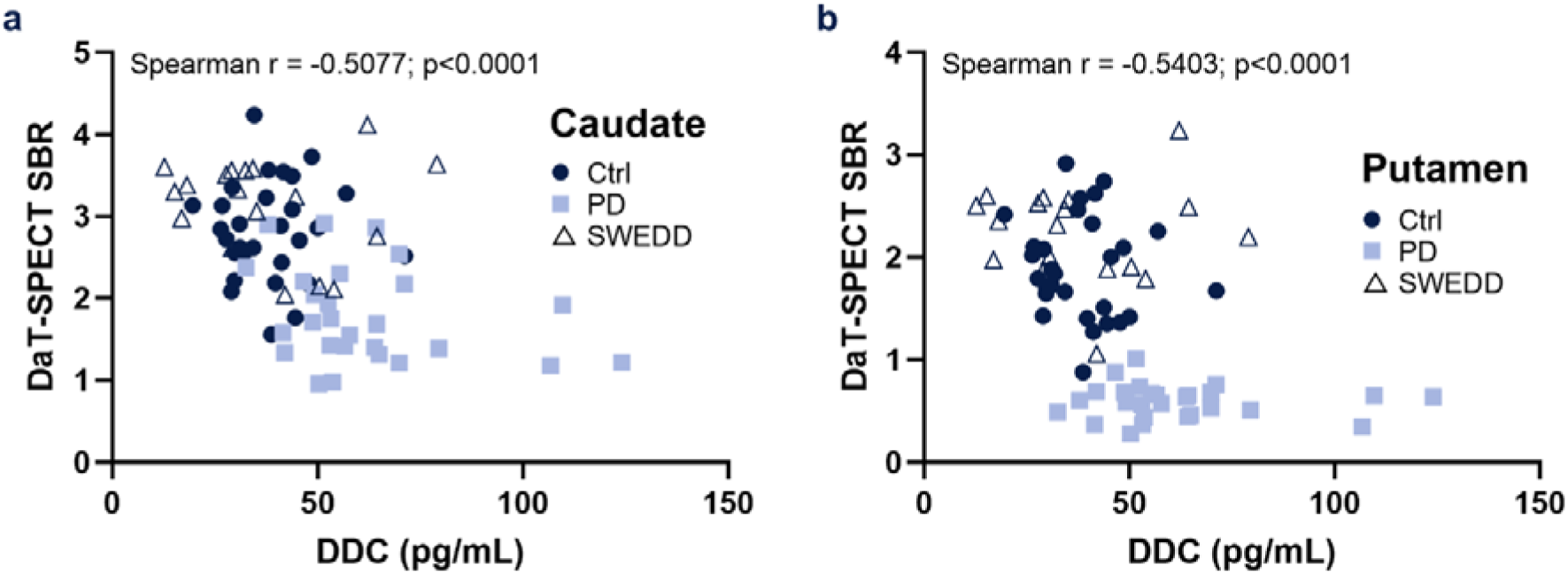
Correlation of CSF DDC levels with DaT-SPECT SBR from the contralateral caudate (**a**) and putamen (**b**).

### Prognostic potential of CSF DDC levels in dopaminergic drug-naïve PD patients

To gain insight into the potential of CSF DDC levels as a prognostic biomarker in PD patients, MDS-UPDRS total score OFF changes over time were compared in patients with low and high baseline DDC levels after splitting at the median. The increase of MDS-UPDRS total scores OFF over time was more pronounced in PD patients with high baseline CSF DDC levels (Figure 4a). In addition, the correlation of baseline CSF DDC levels with MDS-UPDRS total score OFF changes for year 1 through year 8 was evaluated. A significant correlation was first detected at year 5 and persisted through year 8, suggesting DDC may have promise as a prognostic biomarker (Figure 4b).

**Figure 4.**
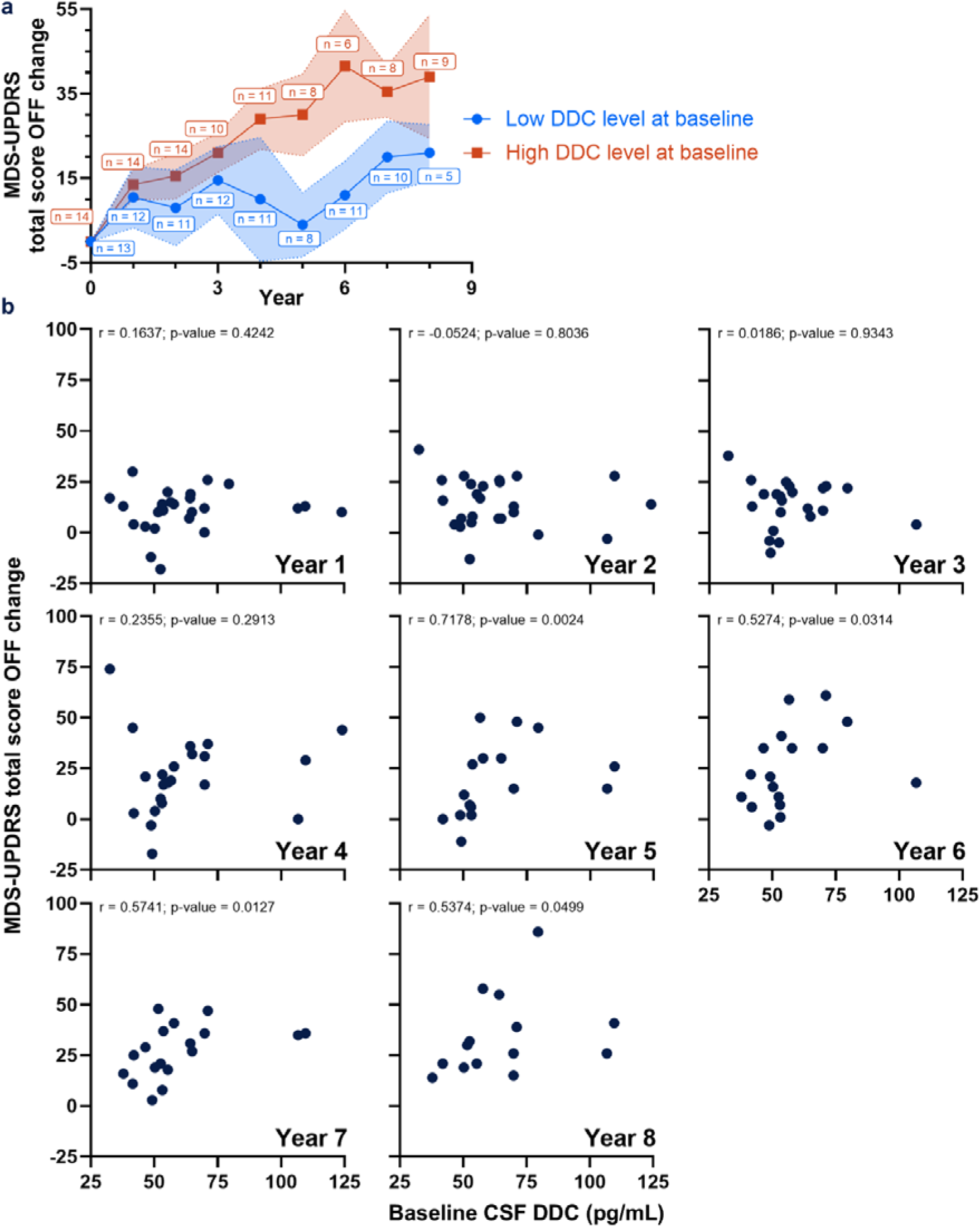
Prognostic potential of CSF DDC levels in dopaminergic drug-naïve PD patients. **a.** MDS-UPDRS total score OFF changes over time in PD patients with low and high baseline CSF DDC levels after splitting at the median. **b.** Spearman correlation of baseline CSF DDC levels with MDS-UPDRS total score OFF changes for years 1–8.

The correlation of baseline CSF DDC levels with MDS-UPDRS part I, MDS-UPDRS part II and MDS-UPDRS part III OFF sub score changes at years 1 through 8 post diagnosis was also investigated. Baseline CSF DDC levels correlated significantly with MDS-UPDRS part II scores at years 5, 6, and 8 as well as with MDS-UPDRS part III scores OFF at years 5 through 7 (Supplementary Figure S6).

### Changes in CSF DDC levels with PD progression and their correlation with LEDD

At the 3-year follow-up visit, the CSF DDC levels were 2.17-fold higher in patients with PD in comparison to Ctrl (p<0.0001). While DDC levels in the Ctrl group did not change from baseline to the 3-year follow-up (p = 0.3304), DDC levels in the PD group increased 1.42-fold (p<0.0001) (Supplementary Figure 7a). In addition, at the 3-year follow-up, CSF DDC levels were significantly higher (1.57-fold) in males than in females in the PD group (p = 0.0145), while there was no significant sex difference in DDC levels among controls (Supplementary Figure S7b).

However, LEDD levels were also significantly higher in males than in females (p = 0.0223) (Supplementary Figure S7c). The relationship between CSF DDC levels and the LEDD was assessed at the 3-year follow-up, and a significant correlation was detected (Spearman r = 0.45, p = 0.0198) (Supplementary Figure S7d). The correlation was stronger in females than in males (Supplementary Figure S7d).

## Discussion

In the current study, we developed and validated a novel ultrasensitive immunoassay for quantification of DDC levels in human CSF. The assay discriminated dopaminergic drug-naïve PD patients from Ctrl and patients with SWEDD with high sensitivity and specificity. At baseline, DDC levels in human CSF were inversely correlated with DaT-SPECT SBR when all study participants were included for analysis. In addition, the study suggested that DDC has potential as a prognostic biomarker in patients with PD. With disease progression, CSF DDC levels increased further but were positively correlated with the LEDD.

Previous investigations have relied on proteomic technologies to determine DDC levels in human CSF [12–16]. However, these approaches are currently limited to relative quantification and associated with high costs. Different proteomics technologies for detection of DDC in CSF have shown differences in diagnostic performance [14] highlighting that DDC detection reagents need to be optimized towards the most relevant DDC proteoforms in biofluids. Upon extensive screening of antibody combinations (Supplementary Figure S2), we developed and validated a DDC immunoassay using highly sensitive SMC technology [25]. Our assay is based on commercially available antibodies and protein calibrator, rendering it accessible to other researchers in the field. Its AUC in a ROC analysis and its sensitivity and specificity for discriminating dopaminergic drug-naïve PD patients from Ctrl were comparable to those described previously for drug-naïve PD and Lewy body disorders using the Olink Explore assay [12,14,16]. Notably, our DDC SMC assay could discriminate dopaminergic drug-naïve patients with PD not only from Ctrl but also from dopaminergic drug-naïve patients with SWEDD. To our knowledge, this is the first study showing that CSF DDC could be used for differential diagnosis of patients with PD versus SWEDD, which are commonly misdiagnosed based on clinical assessment [26–28].

DaT-SPECT imaging of the dopamine transporter can detect dopaminergic deficit in patient brain and facilitate the differential diagnosis of nigrostriatal neurodegenerative Parkinsonian syndromes and parkinsonism that is not caused by nigrostriatal neurodegeneration [4,29]. In the current study, an inverse correlation was detected between CSF DDC levels and DaT-SPECT SBR at baseline when analyzing all groups combined. These data, in parallel with unchanged CSF DDC levels in patients with SWEDD, support the hypothesis that the elevation of CSF DDC levels in PD patients is connected to dopaminergic deficit. However, no significant correlation was observed between CSF DDC levels and DaT-SPECT SBR in the PD group only. This observation agrees with a previous study on dopaminergic treatment-naïve PD patients from two different cohorts [17]. It has been hypothesized that the elevation of CSF DDC may be the consequence of a compensatory reaction to dopaminergic cell loss [12–14,16,17]. However, the spatial and temporal relationship between these two processes is not known. Studies employing longitudinal DaT-SPECT and fluorodopa positron emission tomography as well as CSF sampling could help inform on the relationship between dopaminergic cell loss and DDC levels. Moreover, it would be interesting to investigate the potential of CSF DDC for other dopaminergic deficiencies, including restless legs syndrome, depression, schizophrenia, and attention deficit hyperactivity disorder [30–33].

CSF DDC levels in patients with PD were further elevated at the three-year follow-up visit. The reasons underlying the increase of DDC in CSF of patients with PD have not been definitively elucidated. Several potential mechanisms, including the result of neuronal loss, a compensatory reaction to degeneration of dopaminergic neurons, and a possible dopaminergic treatment effect, have been discussed [12,15,17]. Our current study was not designed to elucidate mechanistic questions. Nevertheless, we observed a weak correlation between CSF DDC levels in PD patients three years post diagnosis and the LEDD. This finding aligns with other studies reporting weak positive correlation or no significant correlation [12,13,17]. In contrast to CSF, elevated DDC levels in plasma are strongly driven by dopaminergic treatment [18]. Since DDC levels in plasma are much higher than in CSF (approximately 150-fold using our assay; data not shown) and proteins cross the blood-CSF barrier to a small extent [34], it is possible that DDC levels in CSF are at least partially influenced by increased levels in plasma. However, an interventional study with biofluid sampling before and after onset of dopaminergic treatment may be required to elucidate to what extent it influences DDC levels.

While at baseline there were no significant differences in CSF DDC levels between males and females in any of the groups in the current study, at three years post diagnosis there was a 1.56-fold increase of CSF DDC levels in male compared to female patients with PD (p = 0.0145). Even though the underlying mechanism has not been elucidated, sex differences in the dysfunction of the dopaminergic system in PD have been reported, with a predisposition to nigrostriatal derangement in males and mesolimbic system changes in females test [35]. Moreover, males had a higher LEDD than females which could be a confounding factor (Supplementary Figure S7d).

Most interestingly, PD patients with high baseline CSF DDC levels were more likely to have a higher change in MDS-UPDRS total score (OFF) with disease progression than PD patients with low baseline CSF DDC levels (Figure 4a). Baseline CSF DDC levels were correlated with changes in MDS-UPDRS total score (OFF) at five to eight years post diagnosis. An analysis of MDS-UPDRS part II and III (OFF) revealed a correlation of CSF DDC levels with motor symptoms. These data suggest that DDC may have potential as a prognostic biomarker for the progression of motor symptoms in patients with PD. However, the sample size was relatively small, and further investigations in larger cohorts are needed to assess the usefulness of DDC in this context.

The current study focused on DDC levels in CSF, which is in direct contact with the central nervous system and thus the most proximal source of biomarkers for neurodegenerative diseases. However, more easily accessible biofluids like plasma are of high interest. Recent studies using the Olink Explore DDC assay showed that plasma DDC is not elevated in dopaminergic drug-naïve patients with PD [12,14,18]. Although our DDC SMC assay correlated moderately with Olink data for CSF samples (Supplementary Figure S5), it is unclear whether the same is true for plasma. Since it is possible that different assays detect different proteoforms of DDC, it would be highly interesting to test our new DDC SMC assay in plasma samples from drug-naïve patients with PD.

This study has several strengths. First, a new immunoassay for quantification of DDC that can be readily used by other researchers was validated and employed. Second, analysis of the well-characterized PPMI cohort enabled us to investigate association of DDC levels in CSF with dopaminergic treatment, functional assessments, and DaT-SPECT imaging. Third, our study included for the first time patients with SWEDD.

However, we acknowledge that our study also has certain limitations. First, the analyzed cohort did not allow to distinguish between changes in CSF DDC levels caused by disease progression versus dopaminergic treatment. Second, the sample size of the investigated cohorts was relatively small, especially for addressing the potential prognostic value of CSF DDC.

## Conclusions

In conclusion, our study on CSF DDC using a novel SMC assay highlighted its potential as diagnostic biomarker for PD and as prognostic biomarker for progression of motor symptoms. Our findings that CSF DDC levels inversely correlate with DaT-SPECT SBR and are unchanged in patients with SWEDD support the hypothesis that elevated CSF DDC levels are connected to dopaminergic deficit in the brain.

## Abbreviations

AD: Alzheimer’s disease
ANOVA: Analysis of variance
AUC: Area under the curve
CSF: Cerebrospinal fluid
Ctrl: Healthy control
CV: Coefficient of variability
DaT: Dopamine transporter
DDC: DOPA decarboxylase
hCSF: Human CSF
L-DOPA: Levodopa
LEDD: L-DOPA equivalent daily dose
LLOQ: Lower limit of quantitation
MDS-UPRDS: Movement Disorder Society Unified Parkinson’s Disease Rating Scale
MSD: Mesoscale discovery
NPX: Normalized protein expression
PBS: Phosphate buffer saline
PD: Parkinson’s disease
PPMI: Parkinson’s Progression Markers Initiative
rhDDC: Recombinant human DDC
ROC: Receiver operating characteristic
S/N: Signal-to-noise
SAAs: Seed amplification assays
SBR: Striatal binding ratio
SD: Standard deviation
SDS-PAGE: Sodium dodecyl sulfate polyacrylamide gel electrophoresis
SMC: Single molecule counting
SPECT: Single-photon emission computed tomography
SWEDD: Scans without evidence for dopaminergic deficit
ULOQ: Upper limit of quantitation

## Supplementary Information

Additional file 1.

## Declarations

### Ethics approval and consent to participate

Details on sample collection were provided by the vendors PrecisionMed Inc. (Solana Beach, CA, USA). The PPMI study (NCT01141023) was approved by the Institutional Review Boards of each PPMI site. Informed written consent was obtained from all subjects at each site.

### Consent for publication

Not applicable

### Availability of data and materials

All data generated and analyzed during the current study are included in this published article and its supplementary file. CSF DDC measurements are available in PPMI database (project 274).

### Authors’ contributions

HA and RGH conceptualized the study. HA designed, performed and analyzed experiments. HA and JEM performed statistical analysis. HA, JEM, SG, SB and RGH interpreted data. HA and RGH obtained human sample cohort. HA and RGH wrote the final manuscript draft. All authors critically reviewed, provided feedback, and approved the final manuscript.

## Acknowledgements

We thank the participants for their generous donation of samples. We thank Dr. Zoya Marinova (ZMedBio, LLC) for manuscript preparation and Dr. Samantha Hutten for her assistance. Data used in the preparation of this article was obtained on (2024-07-29) from the Parkinson’s Progression Markers Initiative (PPMI) database (www.ppmi-info.org/access-data-specimens/download-data), RRID:SCR_006431. For up-to-date information on the study, visit www.ppmi-info.org.

