## Additional file 1 for "A monoclonal antibody-based immunoassay reinforces DOPA decarboxylase in cerebrospinal fluid as a diagnostic and prognostic biomarker for Parkinson’s disease"

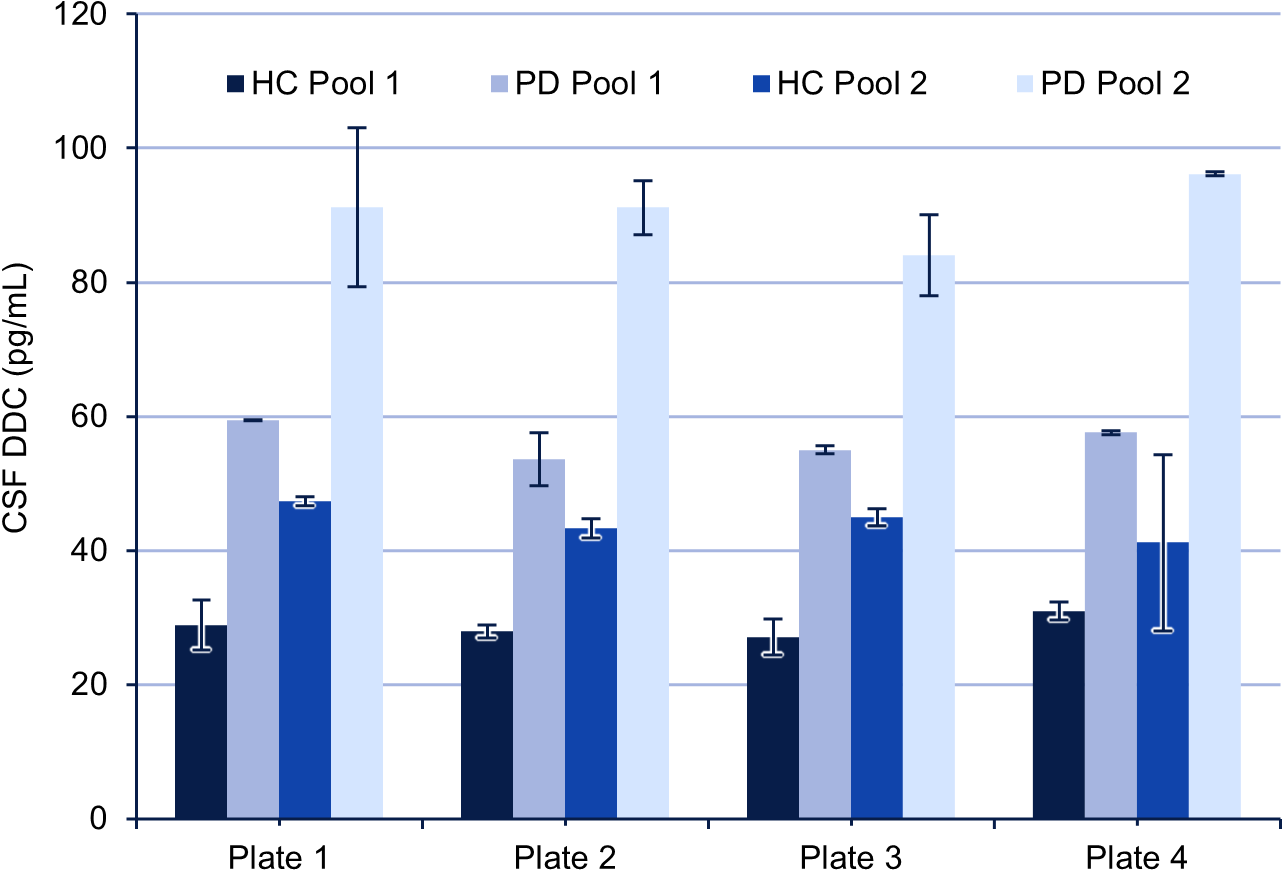

**Supplementary Figure S1.** Quality controls of the DDC assay in the PPMI cohort. Mean ± SD of technical duplicates are plotted.

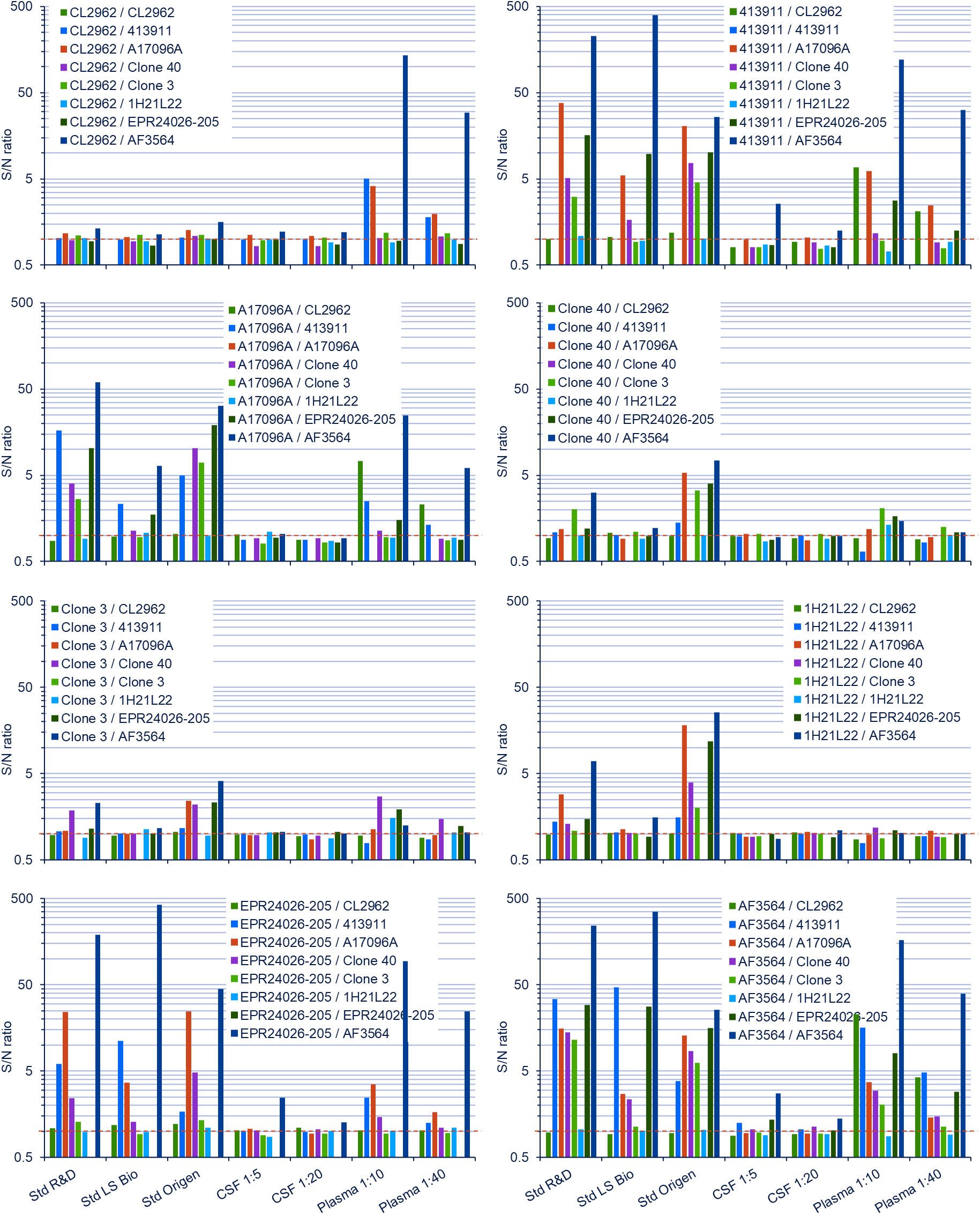

**Supplementary Figure S2.** Screening of antibody combinations using the MSD platform. Signal-to-noise (S/N) ratio of singlicates are plotted.

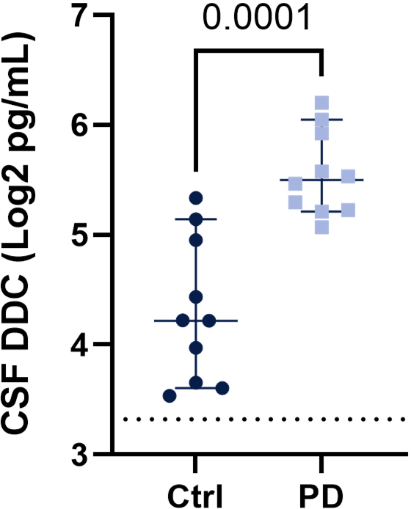

**Supplementary Figure S3.** CSF DDC quantification by SMC assay in the human pilot cohort. For each individual, the mean value of technical duplicates is plotted. Horizontal bars correspond to median ± 95% CI and dashed line to LLOQ. P-value from Welch’s t test on log2 values is indicated.

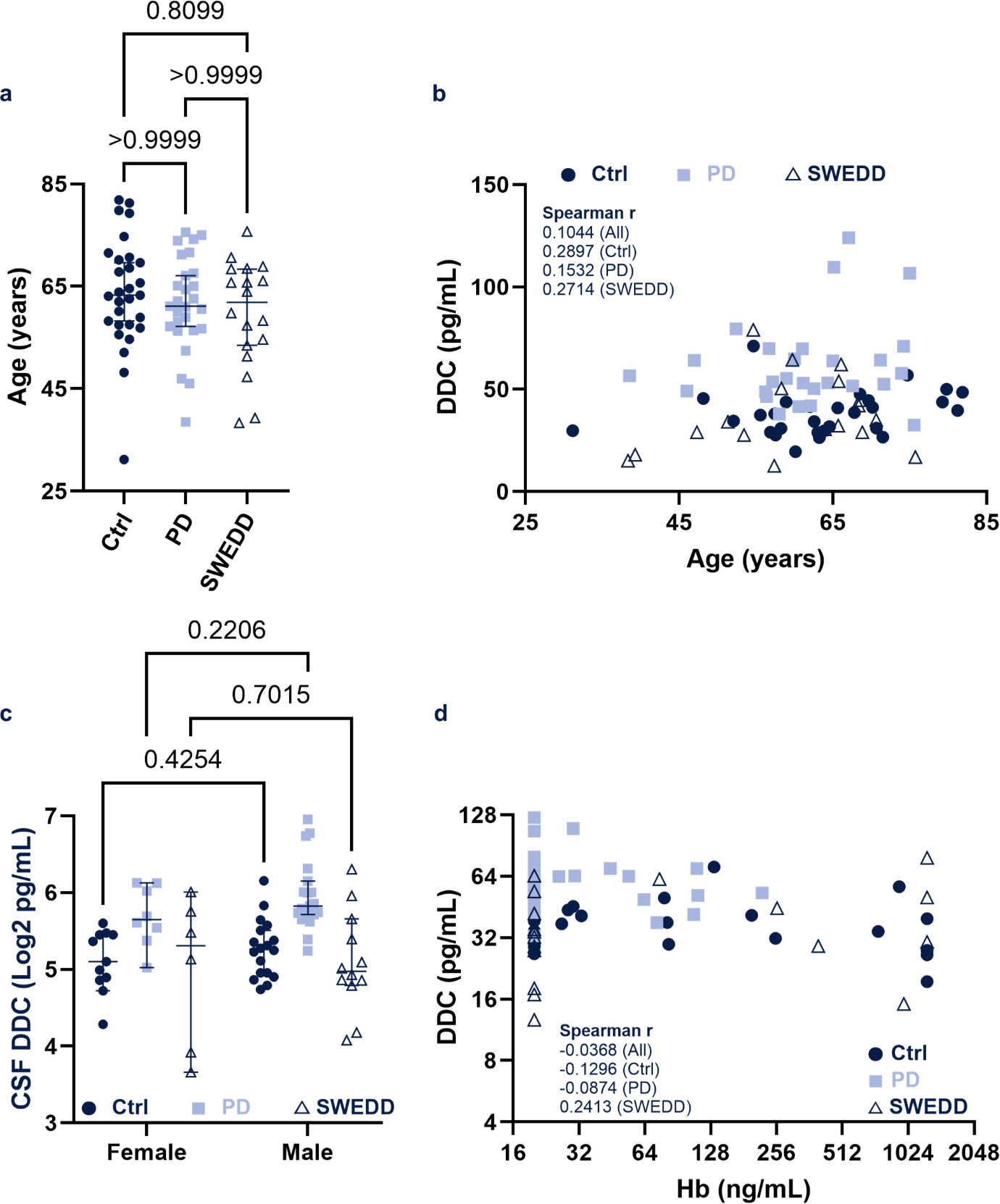

**Supplementary Figure S4.** Relationship of CSF DDC levels with age (**a, b**), sex (**c**), and hemoglobin (Hb) levels **(d)**. For each individual, the mean value of technical duplicates is plotted. Horizontal bars correspond to median ± 95% CI. P-values from Kruskal-Wallis test and Dunn’s multiple comparison (**a**) or ordinary two-way ANOVA and Tukey’s multiple comparison (**c**) is indicated. Spearman correlation r are indicated in (**b**, **c**).

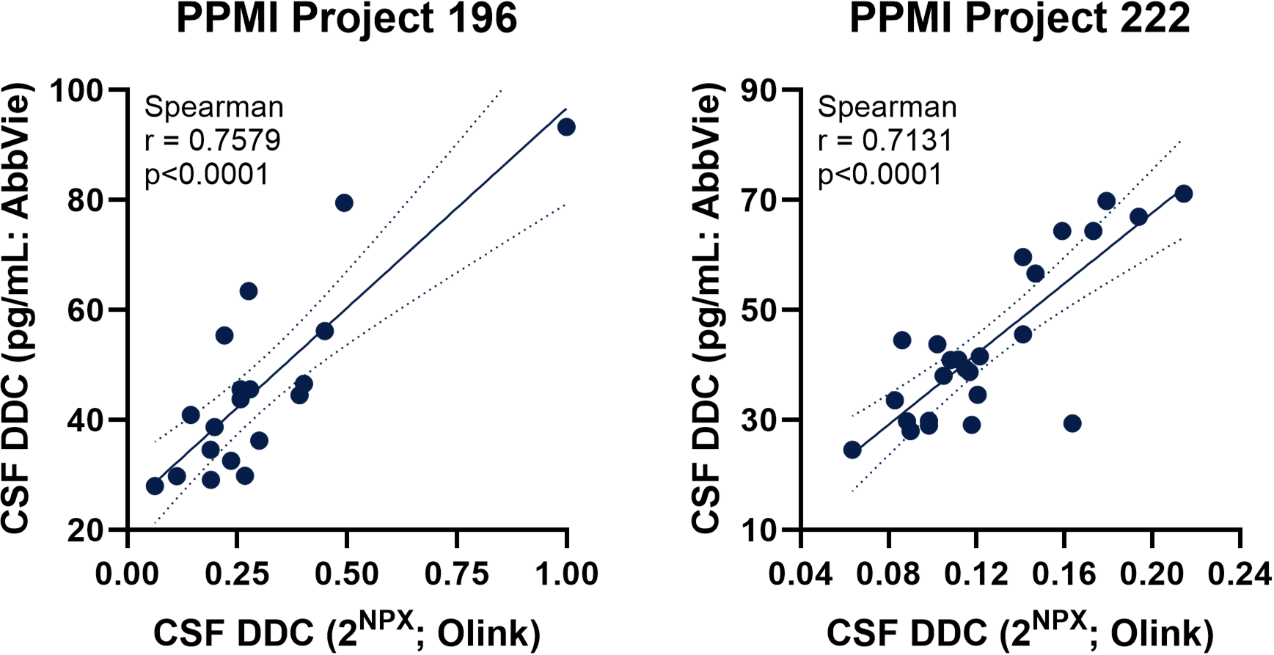

**Supplementary Figure S5.** Spearman correlation between CSF DDC levels determined with the Olink Explore assay in two different PPMI studies and the SMC assay developed in the current study. Linear regression (line) and 95% confidence intervals (dotted lines) are shown.

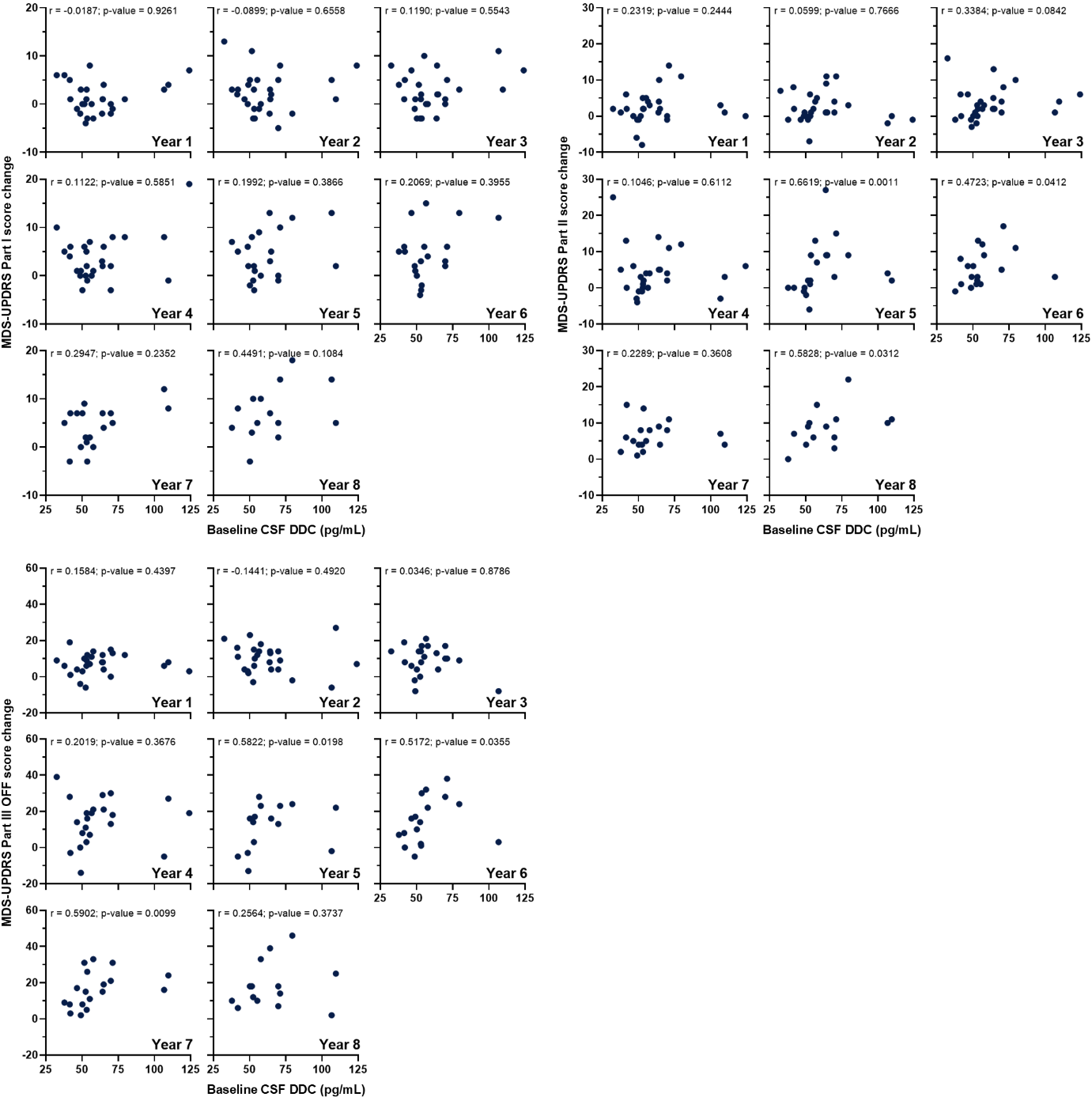

**Supplementary Figure S6.** Spearman correlation of baseline DDC levels with changes in MDS-UPDRS subscores (Part III OFF) from baseline to years 1 through 8 post-PD diagnosis.

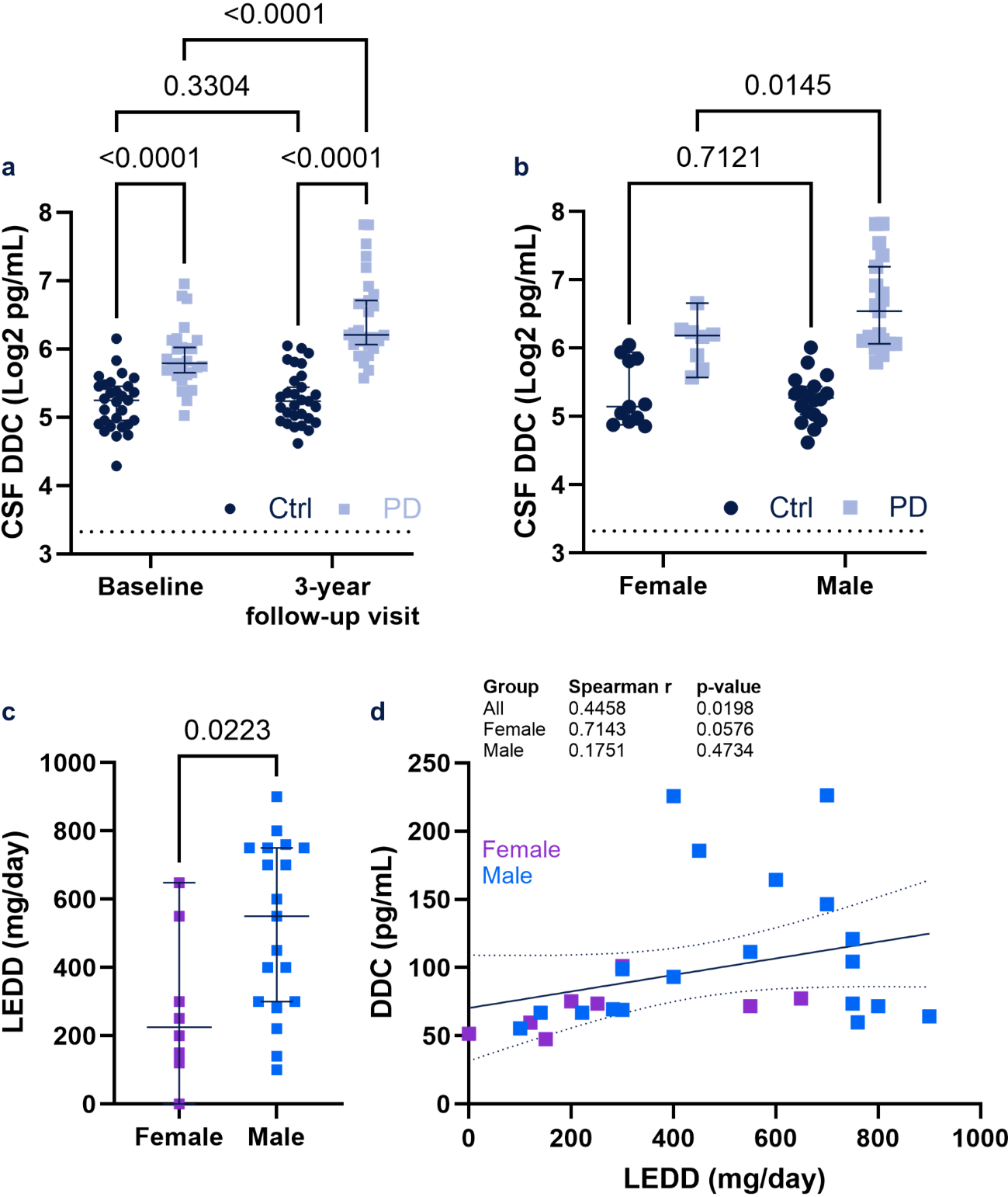

**Supplementary Figure S7.** Changes in CSF DDC levels 3 years after PD diagnosis. **a**. Comparison of CSF DDC levels between PD patients and Ctrl. **b.** Sex difference in DDC concentration. For each individual, the mean value of technical duplicates is plotted. P-values from two-way repeated measures ANOVA (**a**) or ordinary two-way ANOVA (**b**) and uncorrected Fisher’s least significant difference with a single pooled variance is indicated. **c**. Sex difference in LEDD. P-value from Mann-Whitney test is indicated. Horizontal bars correspond to median ± 95% CI. **d.** Spearman correlation between DDC levels and the LEDD.

|  | Ctrl (n = 10) | PD (n = 10) |
| --- | --- | --- |
| **Age (years)** |  |  |
| Mean (SD) | 64.8 (8.1) | 65.6 (8.3) |
| Range | 50-76 | 50-79 |
| **Sex (Number, %)** |  |  |
| Male | 6 (60) | 5 (50) |
| Female | 4 (40) | 5 (50) |

**Supplementary Table S1.** Demographic characteristics of the human pilot cohort. Ctrl: healthy control; PD: Parkinson’s disease; SD: standard deviation

| **Clone** | **Supplier** | **Catalog #** | **RRID** |
| --- | --- | --- | --- |
| CL2962 | Thermo Fisher Scientific | MA5-31379 | AB_2787016 |
| 413911 | R&D Systems | MAB3564 | AB_2088967 |
| A17096A | BioLegend | 869502 | AB_2814636 |
| Clone 40 | Thermo Fisher Scientific | MA5-30483 | AB_2786208 |
| Clone 003 | Thermo Fisher Scientific | MA5-30482 | AB_2786207 |
| 1H21L22 | Thermo Fisher Scientific | 703686 | AB_2848234 |
| EPR24026-205 | Abcam | ab307814 | n/a |
| Polyclonal | R&D Systems | AF3564 | AB_621958 |

**Supplementary Table S2.** List of screened antibodies.

| **Expression System** | **Supplier** | **Catalog #** |
| --- | --- | --- |
| IPLB-Sf21-AE | R&D Systems | 3564-DC-010 |
| Escherichia coli | LifeSpan BioSciences | LS-G489-20 |
| HEK 293 | OriGene Technologies | TP319037 |

**Supplementary Table S3.** List of screened recombinant proteins.
